# Effectiveness of different booster regimens for preventing infection and adverse outcomes in Puerto Rico

**DOI:** 10.1101/2021.12.19.21268070

**Authors:** M.M. Robles-Fontán, R.A. Irizarry

## Abstract

Recent laboratory and observational studies have demonstrated that the COVID-19 vaccine effectiveness wanes over time. In response, several jurisdictions have authorized the administration of booster doses. Since August 13, 2021, Puerto Rico has administered 540,140 booster shots. We used data collected and made public by the Puerto Rico Department of Health (PRDH) to evaluate the effectiveness of four different booster regimens at preventing SARS-CoV-2 laboratory confirmed infections and adverse COVID-19 outcomes. Specifically, we analyzed data from all 115,995 SARS-CoV-2 infections occurring since the vaccination process commenced on December 15, 2020. We combined vaccination status, SARS-CoV-2 test results, and COVID-19 hospitalizations and deaths data, and fit a statistical model that adjusted for time-varying incidence rates and age group, to estimate time-varying vaccine effectiveness against infection and adverse outcomes. We find that, after 6 months, the mRNA-1273 and BNT162b2 effectiveness against infection wanes substantially to 61% (58%-63%) and 36% (34%-39%), respectively, while the Ad26.COV2.S wanes to 35% (31%-39%) after two months. However, after a booster shot of the corresponding initial vaccine manufacturer, effectiveness increased to 87% (83%-91%) and 82% (79%-85%) for mRNA-1273 and BNT162b2, respectively. The effectiveness for Ad26.COV2.S followed by either a mRNA-1273 or BNT162b2 booster increased to 88% (71%-100%), substantially higher than 65% (59%-70%), the peak effectiveness reached with just one shot. We also found that heterologous booster regimens restored effectiveness. Furthermore, we did not observe waning after two months of the booster shot. Finally, we found that all booster regimens provided increased protection against COVID-19 hospitalizations and deaths. Code and data to reproduce the analyses are provided here: https://github.com/rafalab/booster-eff-pr.

## Introduction

The mRNA-1273, BNT162b2, and Ad26.COV2.S vaccines were shown to be highly effective at preventing COVID-19 outcomes in large randomized clinical trials.^1 2 3^ However, several observational studies have demonstrated that vaccine effectiveness wanes over time. A retrospective cohort study that evaluated electronic health records of individuals that are part of the Kaiser Permanente Southern California health-care organization found that BNT162b2 effectiveness against SARS-CoV-2 infections declined from 88% (95% CI 86% -89%) during the first month after completion of primary series to 47% (95 CI 43 -51) after five months.^4^ Two other observational studies found similar results.^5 6^ An observational study, also based on PRDH data, found that effectiveness against laboratory-confirmed SARS-CoV-2 infection, COVID-19 hospitalization and deaths wanes over time for the mRNA-1273, BNT162b2, and Ad26.COV2.S COVID-19 vaccines. At the peak of their protection, these vaccines had an effectiveness of 90% (95 CI 88 - 91), 87% (95 CI 85 - 89), and 58% (95 CI 51 - 65). After four months, effectiveness waned to about 70%, 60%, and 30% for mRNA-1273, BNT162b2, and Ad26.COV2.S, respectively.^7^ Here we analyze a more recent version of these data further to estimate primary series effectiveness past six months and booster regimens effectiveness.

Since the publication of these studies, several jurisdictions have authorized and commenced administering booster shots. An observational study from Israel evaluated vaccine effectiveness for BNT162b2 seven days after receipt of booster dose and compared to those receiving only two doses at least five months before. Booster shot effectiveness was estimated to be 93% (95 CI 88–97) for COVID-19 admission to hospital, 92% (95 CI 82–97) for COVID-19 severe disease, and 81% (95 CI 59–97) for COVID-19 death.^8^ A second observational study from Israel found that a BNT162b2 booster shot administered at least five months after completion of the primary series gave recipients 90% lower COVID-19 mortality in comparison to those who had not received the booster dose. Similarly, the study found that the BNT162b2 booster dose effectiveness against SARS-CoV-2 infection was estimated to be at 83% for those who had received a booster dose in comparison to those who had not.^9^ A third observational study from Israel evaluated BNT162b2 booster shot effectiveness against infection and severe COVID-19 illness across age groups and found no significant difference in effectiveness among age groups.^10^

In Puerto Rico, booster shot administration began on August 13, 2021. By November 28, 2021, at least 2,348,520 of the 3,285,874 individuals living in Puerto Rico had completed the COVID-19 vaccination primary series. Of these, 540,140 individuals have received a booster dose. Furthermore, since the start of the vaccination process to October 15, 2021, Puerto Rico has recorded 115,995 SARS-CoV-2 laboratory confirmed infections, with 32,904 detected after boosters began being administered. We leveraged data collected by the PRDH to estimate time-varying vaccine effectiveness for all three vaccines, mRNA-1273, BNT162b2, and Ad26.COV2.S, and the effect of four different booster regimens.

## Methods

We combined vaccination status, SARS-CoV-2 test results, and COVID-19 hospitalizations and deaths data, which permitted us to quantify the time-varying effectiveness of the mRNA-1273, BNT162b2, and Ad26.COV2.S vaccines past six months and the effect of boosters shots. Specifically, we used data made publicly available by the PRDH.^7^ These data included a table with information for all individuals that have been vaccinated and a table for all individuals with a laboratory-confirmed SARS-CoV-2 infection recorded after December 15, 2020. Both tables included each individual’s age group and gender. The vaccination table also included, for each individual, the date and manufacturer of each administered vaccine dose. This information was also included for vaccinated individuals represented in the cases table. The cases table also denoted if each individual had been hospitalized or died. Since in November there was a major outbreak in prisons, and prisoners were vaccinated almost exclusively with the BNT162b2 vaccine, we removed these individuals from the analysis to avoid bias. The results with and without these individuals were similar, and because the code and data are available, one can easily run both analyses.

From the vaccine data, we were able to compute daily counts of unvaccinated individuals, individuals receiving their first doses, individuals becoming fully vaccinated, and individuals receiving a booster shot for each vaccine manufacturer and demographic group. Furthermore, we computed counts of individuals vaccinated on day *t*_*v*_ and counts of individuals boosted on day *t*_*b*_ for each pair of dates *t*_*v*_ *< t*_*b*_, for each demographic group and pair of vaccine manufacturers.

From the cases table, we were able to compute daily counts of cases, hospitalizations, and deaths among the unvaccinated, the vaccinated without booster, and the vaccinated with booster for each demographic group. For the vaccinated, we further divided the counts by manufacturer combination. For the vaccinated, we also calculated the number of days since they were vaccinated, and for those that received a booster, the time since they received a booster. We computed rates for unvaccinated individuals and defined the population as the number of individuals that were unvaccinated and had not been infected within the last three months.

With these data in place, we were able to apply a previously developed statistical approach to estimate primary series vaccine effectiveness and booster regimen effectiveness as a function of time.^7^ Note that this is different from estimating effectiveness as a function of calendar date since the date individuals became fully vaccinated vary from January to December.

## Results

### Booster data

As of November 28, 2021, 2,348,520 of the 3,285,874 individuals living in Puerto Rico had been fully vaccinated and 540,140 of these had received the booster shot. The majority of people received booster shots of the same manufacturer as those in their initial series. Few had completed mix-n-match booster regimens. In all, 212,922 mRNA-1273, 322,045 BNT162b2 and 5,173 Ad26.COV2.S booster shots were administered in Puerto Rico (Table 1). Because so few Ad26.COV2.S booster doses had been administered, we removed these from the analysis comparing booster regimens.

**Table 1:** For each combination of initial series and booster manufacturers, we show the number of individuals receiving the respective combination.

| **Initial series** | **Booster** | **Total** |
| --- | --- | --- |
| BNT162b2 | BNT162b2 | 301,443 |
| mRNA-1273 | mRNA-1273 | 200,041 |
| Ad26.COV2.S | BNT162b2 | 10,719 |
| mRNA-1273 | BNT162b2 | 9,883 |
| Ad26.COV2.S | mRNA-1273 | 9,048 |
| Ad26.COV2.S | Ad26.COV2.S | 5,140 |
| BNT162b2 | mRNA-1273 | 3,833 |
| mRNA-1273 | Ad26.COV2.S | 23 |
| BNT162b2 | Ad26.COV2.S | 10 |

### Vaccination series and booster effectiveness

We estimated time-varying effectiveness of a primary series of COVID-19 vaccines for three COVID-19 outcomes: SARS-CoV-2 laboratory confirmed infection, COVID-19 hospitalization and death. Vaccine effectiveness against SARS-CoV-2 infection after completion of the primary series reached a peak effectiveness right after completion 89% (87%-90%), 85% (84%-87%), and 65% (59%-70%) for mRNA-1273, BNT162b2, and Ad26.COV2.S, respectively. After six months, vaccine effectiveness dropped to 61% (58%-63%), 36% (34%-39%), and 36% (27%-45%) for mRNA-1273, BNT162b2, and Ad26.COV2.S, respectively. However, booster shots restore vaccine effectiveness against infection (Figure 1, Table 2). Specifically, homologous booster regimens restored vaccine effectiveness infection to 87% (83%-91%) for mRNA-1273 + mRNA-1273 and to 82% (79%-85%) for BNT162b2 + BNT162b2. Heterologous booster regimens restored vaccine effectiveness against infection to 88% (71%-100%) for JSN + mRNA-1273 or BNT162b2 and to 81% (56%-100%) for mRNA-1273 + BNT162b2 or BNT162b2 + mRNA-1273.

**Table 2:** Effectiveness against SARS-CoV-2 laboratory confirmed infection with 95% confidence intervals by time period since full vaccination for each vaccine manufacturer.

| **Status** | **mRNA-1273** | **BNT162b2** | **Ad26.COV2.S** |
| --- | --- | --- | --- |
| Right after vaccination | 89% (87%-90%) | 85% (84%-87%) | 65% (59%-70%) |
| 0 to 2 months after vaccination | 85% (84%-86%) | 80% (79%-81%) | 59% (55%-62%) |
| 2 to 4 months after vaccination | 75% (74%-76%) | 62% (61%-63%) | 35% (31%-39%) |
| 4 to 6 months after vaccination | 68% (66%-69%) | 58% (56%-59%) | 38% (34%-42%) |
| More than 6 months after vaccination | 61% (58%-63%) | 36% (34%-39%) | 36% (27%-45%) |
| Boosted | 87% (83%- 91%) | 82% (79%- 85%) | 88% (71%-100%) |

**Figure 1:**
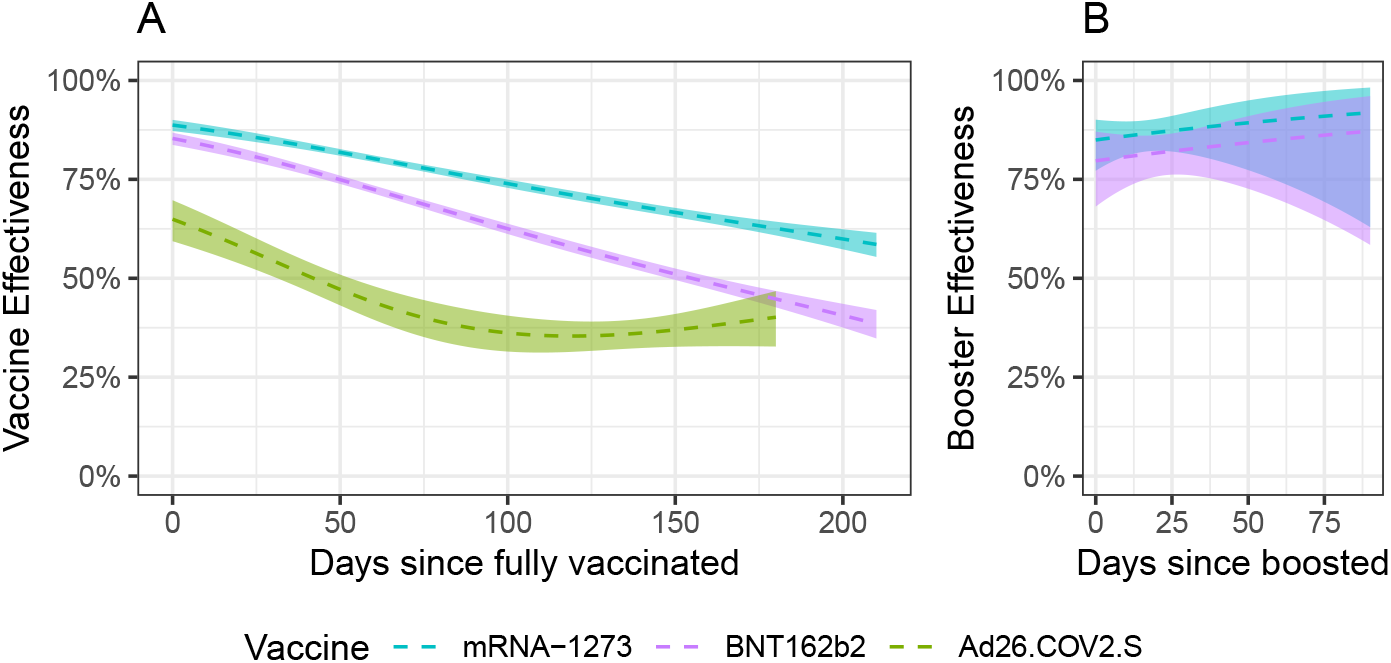
Estimated effectiveness against infection plotted against days since the individuals were fully vaccinated. The ribbons represent point-wise 95% confidence intervals. A) Vaccine effectiveness against infections by manufacturer for the entire study period. B) Booster effectiveness against infections by manufacturer.

Furthermore, we estimated vaccine effectiveness against COVID-19 hospitalization and death. During the first six months after completion of the primary series, vaccine effectiveness against COVID-19 hospitalization averaged 95% (92%-97%), and 94% (91%-96%), for mRNA-1273, BNT162b2, respectively. After six months, effectiveness decreased to 81% (75%-86%) and 79% (74%-84%) for mRNA-1273 and BNT162b2, respectively. With the Ad26.COV2.S, effectiveness averaged 77% (61%-86%) for the first two months and dropped to 66% (59%-72%) after that (Figure 2, Table 3). During the first six months after the completion of the primary series, vaccine effectiveness against COVID-19 deaths averaged 99% (96%-100%), and 99% (97%-100%), for mRNA-1273, BNT162b2, respectively. After six months, effectiveness decreased to 88% (81%-95%) and 95% (91%-98%) for mRNA-1273 and BNT162b2, respectively. For Ad26.COV2.S, effectiveness averaged 74% (32%-90%) for the first two months and dropped to 69% (58%-80%) after that (Figure 2, Table 3). Due to small sample size, to increase statistical power, for hospitalizations and deaths, we computed vaccine effectiveness for all individuals receiving a booster shot. We found that a booster dose restored effectiveness to 89% (82%-96%) against hospitalization and 94% (84%-100%) against death (Table 4).

**Table 3:** Effectiveness against COVID-19 adverse events with 95% point-wise confidence intervals by time period after completion of dose. Boosters are recommended after 6 months for mRNA-1273 and mRNA-1273 and after two months for Ad26.COV2.S.

| **Outcome** | **Time period** | **mRNA-1273** | **BNT162b2** | **Ad26.COV2.S** |
| --- | --- | --- | --- | --- |
| Hospitalization | Right after dose completion | 95% (92%- 97%) | 94% (91%- 96%) | 77% (61%- 86%) |
| Hospitalization | Not eligible for booster | 93% (92%- 94%) | 88% (87%- 89%) | 80% (73%- 87%) |
| Hospitalization | Booster recommended | 81% (75%- 86%) | 79% (74%- 84%) | 66% (59%- 72%) |
| Deaths | Right after dose completion | 99% (96%-100%) | 99% (97%-100%) | 74% (32%- 90%) |
| Deaths | Not eligible for booster | 94% (93%- 96%) | 92% (91%- 94%) | 79% (62%- 92%) |
| Deaths | Booster recommended | 88% (81%- 95%) | 95% (91%- 98%) | 69% (58%- 80%) |

**Table 4:** Booster effectiveness against COVID-19 hospitalization and death.

| **COVID-19 Outcome** | **Booster effectiveness** |
| --- | --- |
| Hospitalization | 89% (82%- 96%) |
| Death | 94% (84%-100%) |

**Figure 2:**
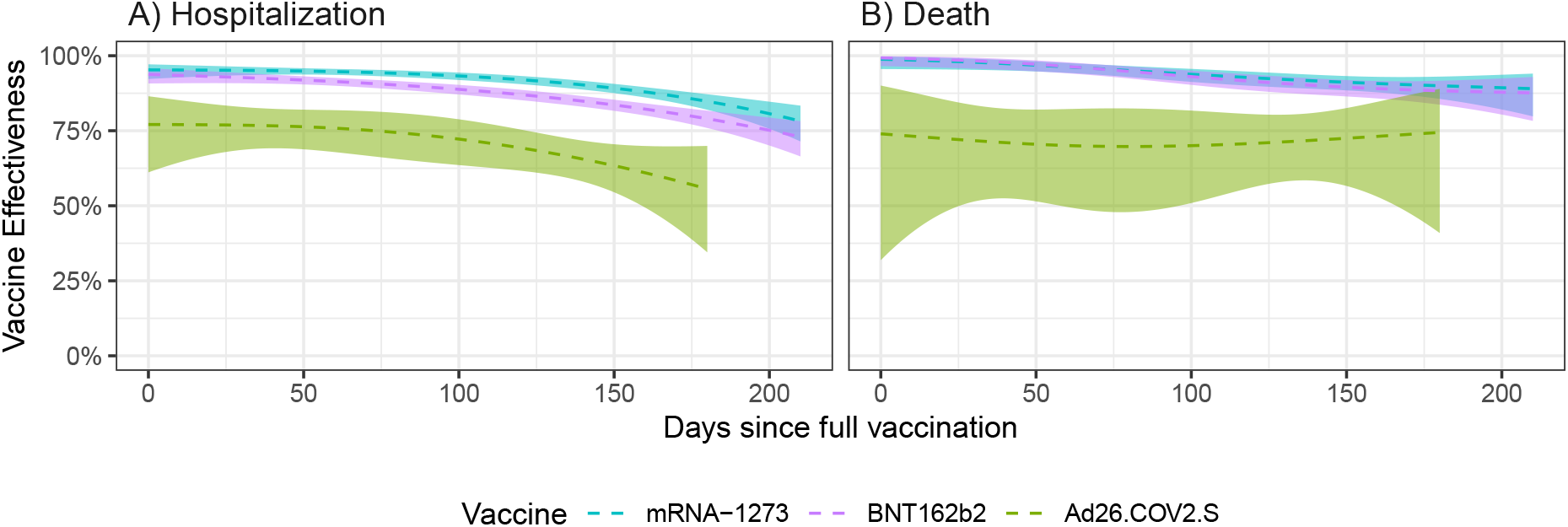
Estimated effectiveness against SARS-CoV-2 laboratory confirmed infection plotted against days since the individuals were fully vaccinated. The ribbons represent point-wise 95% confidence intervals. A) Hospitalization. B) Death.

## Discussion

In this observational study, we found that primary series vaccine effectiveness against all COVID-19 outcomes decreases substantially over time. However, COVID-19 mRNA-1273 and BNT162b2 booster regimens restore effectiveness for all three vaccine primary series against SARS-CoV-2 laboratory confirmed infections. In the case of Ad26.COV2.S, the booster regimen effectiveness againts SARS-CoV-2 laboratory confirmed infection improves substantially over the primary dose effectiveness. Similarly, booster regimens restore effectiveness against COVID-19 hospitalizations and deaths. Booster vaccine effectiveness against SARS-CoV-2 laboratory confirmed infection remains constant over a period of three months. These findings highlight the importance of booster shots to prevent adverse COVID-19 outcomes and minimize stress on health care systems.

## Data Availability

All data produced are available online at https://github.com/rafalab/booster-eff-pr

## Acknowledgments

We thank Dr. Iris Cardona, PRDH Chief Medical Officer, and Dr. Carlos Mellado, Puerto Rico’s Secretary of Health, for their continued support and collaboration. We also thank Elvis Nieves, BioPortal Coordinator, and his team for their help and collaboration.

## Notes

### Competing Interest Statement

The authors have declared no competing interest.

### Funding Statement

This study was partially funded by NIH grant R35GM131802

### Author Declarations

All data and code necessary to reproduce the analysis are here: https://github.com/rafalab/booster-eff-pr

